# Neurocysticercosis in California: Clinical Characteristics and Associated Comorbidities Across the University of California Health System

**DOI:** 10.64898/2026.09.02.26362030

**Authors:** Tayf Mohammed Islam, Samantha Eve Allen

## Abstract

**Introduction:** Neurocysticercosis (NCC) is an infection of the central nervous system caused by the tapeworm Taenia solium. Despite documented increases of NCC in California, large-scale studies characterizing affected populations remain limited. The spectrum of NCC associated neuropsychiatric comorbidities has not been well defined in U.S. populations. This study aimed to estimate the prevalence of NCC within the University of California (UC) Health System, characterize the population demographic and clinical features, and identify factors associated with epilepsy among individuals with NCC.

**Methodology/Principal Findings:** We conducted a cross-sectional analysis of electronic health records from 2012 through March 2026 using the UC Health Data Warehouse, which integrates data from six UC Health systems. Among individuals aged ≥ 1 year, there were 713 individuals with a diagnosis of NCC corresponding to a prevalence of 6.17 per 100,000. A comparison group was randomly selected at a 1:10 ratio. Relative to the comparison group, NCC patients were significantly older, more likely to identify as Hispanic or Latino, more likely to speak Spanish as a primary language, and more likely to have Medicaid insurance. NCC was strongly associated with epilepsy (OR = 19.20, 95% CI: 15.74–23.46), as well as migraines, cognitive impairment, mood disorders, Parkinsons disease, substance use, stroke, and sleep disorders. After adjusting for age, gender, and ethnicity, male NCC patients had 26% higher odds of epilepsy compared to female patients.

**Conclusion/Significance:** In addition to epilepsy, NCC was associated with a broad range of neuropsychiatric comorbidities and occurred disproportionately among patients more likely to face socioeconomic and structural barriers to care, including limited insurance coverage, language barriers, and immigration-related challenges. These findings highlight the importance of recognizing NCC in non-endemic settings, where this often underrecognized disease contributes substantially to neurologic morbidity in a vulnerable population.

**Author Summary:** Neurocysticercosis is an infection of the brain caused by the larvae of the tapeworm Taenia solium. It is commonly found in regions of Latin America, Africa, and Asia. It is among the leading causes of acquired epilepsy worldwide. Although it is thought to be relatively rare in the United States, we suspected that it may be more common than currently recognized, particularly in states with large immigrant populations.

We reviewed 14 years of medical records from over 11 million patients across 6 hospitals within the University of California Health System to find all individuals diagnosed with neurocysticercosis and compared them with a similar group of patients without it. We found about 6 cases of neurocysticercosis per 100,000 patients. These patients were more likely to have illnesses such as epilepsy, migraines, cognitive impairment, Parkinson’s disease, mood disorders, and strokes compared to individuals without neurocysticercosis. They were also more likely to be Spanish-speaking, Hispanic or Latino, and insured through Medicaid.

Our findings suggest that NCC remains an underrecognized cause of neurological illness in California and highlight disparities associated with the disease, underscoring the need for further investigation of these relationships as well as equity-focused strategies to improve NCC diagnosis and care.

## Introduction

Neurocysticercosis (NCC) is an infection of the central nervous system (CNS) caused by larval cysts of the tapeworm *Taenia solium.*[1–6] It is among the leading causes of acquired epilepsy worldwide and represents an important but underrecognized public health concern in the United States.[1,3,5] The World Health Organization (WHO) recognizes NCC as a “major neglected disease” due to its substantial health burden and its disproportionate impact on under-resourced populations.[4,6] Infection occurs through ingestion of *T. solium* eggs in contaminated food or water. After ingestion, the eggs release larvae that penetrate the intestinal wall and spread to various tissues where they form cysts.[1,2,3,6] The larvae have a strong tropism for the CNS, and when cysts form in the CNS, this infection is considered NCC.[6] While NCC is most common in low to middle-income regions of Latin America, Africa, and Asia where *T. solium* in endemic, NCC cases in the United States remain prevalent among immigrants and travelers to and from these endemic areas.[1–3,5,6] Despite the recognized burden of NCC in the United States, there is no unified national surveillance system for the disease. Reporting requirements vary by state, and many states do not mandate reporting at all,[7,8] limiting the ability to estimate disease burden and conduct large-scale epidemiologic studies nationally. Studies by Croker et al. in 2012 [8] and Sorvillo et al. in 2004 [9] characterized the epidemiology of NCC in California and established its importance as a statewide public health concern. No large-scale studies in California have been conducted since these studies. Updated data are needed to characterize the current demographic and clinical profiles of individuals with NCC to inform public health and clinical practice.

Clinical manifestations of NCC vary widely depending on factors such as the location, size, number, and stage of NCC associated lesions within the CNS, and the host’s immune response.[1–3, 5,6] Many infections may remain asymptomatic, while others present with a range of neurologic symptoms.[1,3,5,6] Seizures, headaches, and migraines are the most well-documented symptoms, however, NCC can present with an even broad spectrum of neurologic manifestations, reflecting the interplay between parasite-related factors (e.g., lesion number, location, size, and stage) and host-related factors (e.g., immune response and preexisting patient characteristics).[2] The full spectrum of neuropsychological conditions associated with NCC remains incompletely characterized.[5]

Epilepsy stands out as one of the most consequential and extensively studied outcomes of NCC. Epilepsy itself is a major neurologic disorder affecting more than 50 million people globally and contributes to a significant physical, psychosocial, and economic burden for those affected and their families.[10] In the United States, these burdens may be further amplified among immigrant populations, who disproportionately face structural barriers to healthcare including gaps in insurance coverage, limited access to specialty care, and language barriers, that may delay diagnosis and limit treatment access.[11,12]

This study utilized the University of California Health Data Warehouse (UCHDW) to characterize the demographic and clinical features of individuals with NCC across the University of California (UC) Health System. The UC Health system consists of UC campuses at Davis, UC San Diego, Los Angeles, San Francisco, Irvine and Riverside. California’s large, diverse patient population, shaped by substantial immigration from *T. solium*-endemic regions of Latin America and Asia,[13] offers a valuable setting to better understand the presentation, comorbidities, and healthcare burden of NCC in a non-endemic region.

This study had three specific aims:

1. To describe the prevalence and clinical and demographic characteristics of UC Health patients with NCC.
2. To evaluate associations between NCC and neurologic and psychiatric comorbidities including epilepsy, migraine/headache, cognitive impairment, mood disorders, schizophrenia, stroke, substance use disorders, and sleep disorders, relative to uninfected patients.
3. To evaluate the independent association between NCC and epilepsy, and to identify demographic and clinical characteristics associated with epilepsy, after adjusting for selected covariates.

On an individual level, understanding the independent association between NCC and epilepsy, along with the demographic and clinical factors that further elevate risk for epilepsy, may support earlier identification and monitoring of patients at highest risk. At a population level, these findings may inform clinical and public health approaches to earlier intervention and prevention of epilepsy-related complications.

## Methods

### Data Source

We utilized de-identified, Health Insurance Portability and Accountability Act (HIPAA)- compliant data from the UCHDW, which integrates electronic health record (EHR) data from the six UC health campuses. Data between 01/01/2012 - 03/31/2026 was available at the UCHDW at the time of the study, which was used as the study period. The UCHDW was developed and maintained by the Center for Data-driven Insights and Innovation (CDI2) and harmonized using the Observational Medical Outcomes Partnership (OMOP) Common Data Model (V5.4), an open-source framework developed by the Observational Health Data Sciences and Informatics (OHDSI) program.[14] OMOP converts data from disparate source systems into a standardized format and vocabulary, allowing for consistent identification of diagnoses, medications, and procedures across institutions. During the study period, the UCHDW contained records for approximately 11,560,754 patients.

### Study Population

Individuals aged ≥ 1 year with a diagnosis of NCC were identified using International Classification of Diseases, Tenth Revision (ICD-10) and Systematized Nomenclature of Medicine (SNOMED) codes (ICD-10: B69 for cysticercosis, B69.0 for cysticercosis of central nervous system (NCC); SNOMED: 59051007 for cysticercosis, 230215006 for cerebral cysticercosis, 187148002 for cysticercosis of central nervous system, 1163537001 for cysticercosis myelitis, and 441460004 for cysticercosis of brain). We utilized OMOP’s hierarchical vocabulary structure to capture related diagnoses. The group of patients identified consisted of patients diagnosed with “cysticercosis,” “cysticercosis of central nervous system,” and “*Taenia solium* infection”. *Taenia solium* infection (SNOMED: 240818) refers to taeniasis, which results from infection with the adult tapeworm, which by itself would not constitute a cysticercosis or NCC diagnosis.[4] Patients with only a *Taenia solium* and/or cysticercosis diagnosis were excluded due to lack of clarity regarding an NCC diagnosis.

A comparison group was generated by randomly selecting 10 individuals per NCC case at the same UC campus with at least 5 total visits to the health system and no diagnosis for *T. solium* infection, cysticercosis, or NCC. A minimum threshold of 5 visits was set to ensure that comparison patients had sufficient contact with the healthcare system for any comorbidities to be reliably documented and to exclude individuals with only brief encounters (e.g. isolated emergency room visits) where such conditions may not have been charted. Stratifying by campus ensured a representative sample from each UC campus and avoided overrepresentation of larger campuses with higher patient populations.

No a priori sample size or power calculation was performed. The number of NCC cases reflects all individuals meeting eligibility criteria within the UCHDW during the study period, and a 1:10 case-to-comparison ratio was selected to increase statistical power for detecting associations, given the relatively small number of available cases.

### Comorbidities and Clinical Variables

Comorbidities among the study population were identified using the same process as NCC. Conditions of interest included migraines and headaches (S1A Table), cognitive impairment (S1B Table), mood disorders (S1C Table), schizophrenia (S1D Table), substance use disorders (S1E Table), stroke (S1F Table), Parkinson’s disease (S1G Table), and sleep disorders (S1H Table), and epilepsy (S1I Table). Additional patient-level variables extracted for each individual included date of first diagnostic inclusion in the chart, sex, race, ethnicity, primary spoken language, insurance type, area deprivation index (ADI), birth year, and death date (if applicable). To protect patient privacy, anyone aged 89+ had a birth year set to 1937 in the UCHDW at the time of the study per UCHDW policy. ADI is a validated neighborhood-level measure of socioeconomic disadvantage that incorporates indicators across four domains: income, education, employment, and housing quality. [15] Higher ADI scores reflect greater levels of socioeconomic disadvantage.[15] Primary spoken languages with small cell counts (less than 10 for any group) including Chinese, Vietnamese, Arabic, Korean, Farsi, and Russian were collapsed into an “other” category. For NCC patients with multiple records for insurance coverage, we selected the insurance coverage they had closest to, but before their NCC diagnosis date. For comparison patients, we selected the first recorded insurance type.

### Study Design Considerations

This study employed a cross-sectional design. Diagnostic dates for NCC were not used to define disease onset, as dates recorded in the EHR reflect when a diagnosis was first documented rather than when the condition truly began. Therefore, the analysis focused on the presence or absence of conditions rather than temporal relationships between them.

### Statistical Analysis

Data extraction was performed using SQL within the Spark analytical platform Databricks, and all statistical analyses were conducted in R (version 4.4.2) [16] using the tidyverse,[17] sparklyr,[18], broom[19], effectsize[20] packages. Descriptive statistics were calculated as frequencies and proportions for categorical variables and means with standard deviations for continuous variables. Continuous variables including age, ADI, and age at death were compared across groups using two-tailed Welch Two Sample T-Test. Comparative analyses of categorical variables between NCC cases and the comparison group were performed using chi-square tests. Fisher’s Exact Test was used in place of chi-square when one or more expected cell counts were less than ten. For larger contingency tables where any cell count was zero or where there was a combination of very small and very large cell counts, a Monte Carlo approximation based on 100,000 simulations was used to estimate *p*-values in place of standard Fisher’s Exact Test. Statistical significance was evaluated using a predetermined alpha (α) level of 0.05. For the purposes of regression analyses, NCC was treated as the primary exposure of interest and comorbidities such as epilepsy as the outcome. Univariate logistic regression models were used to estimate odds ratios (ORs) and 95% confidence intervals (CIs) for all comorbidities. A multivariable logistic regression model was used to identify potential demographic and clinical characteristics associated with epilepsy among the study population.

Covariates for the multivariable logistic regression model were selected a priori based on clinical and epidemiological relevance and data availability, and included age, sex, ethnicity, and ADI.

For the multivariable logistic regression model evaluating factors associated with epilepsy, patients with missing ADI values were excluded from the analysis (19 patients with NCC and 395 comparison patients). Patients whose sex was categorized as “Other” or “Unknown” were also excluded because of small cell counts. Individuals with unknown ethnicity were retained in the analysis due to the relatively large number of observations in this category. The resulting adjusted epilepsy model included 7,422 observations.

Multicollinearity among these covariates was assessed using the condition number of the model matrix. Because age and ADI are measured on substantially different numeric scales, both were centered and standardized prior to this calculation to avoid inflated results driven by scale differences rather than true collinearity between covariates (unscaled κ = 201; after centering and standardizing, κ = 4.07, indicating no substantial collinearity). Continuous covariates were centered and standardized only for the purpose of assessing multicollinearity; the reported model (Table 5) uses covariates in their original units for interpretability. Interaction terms were not formally tested, as the primary aim of the adjusted model was to estimate the independent association between NCC and epilepsy after adjusting for selected covariates rather than to evaluate effect modification.

## Results

### Prevalence of Neurocysticercosis

Within the total UC Health patient population of 11,560,754 individuals during the study period, 713 cases of NCC were identified between January 2012 and March 2026. This corresponded to a prevalence of 6.17 cases per 100,000 patients for NCC.

### Cohort Characteristics

The final study population included 713 individuals with a diagnosis of NCC and 7130 comparison patients without NCC or associated diagnoses, for a total analytic sample of 7843 patients. Demographic and clinical characteristics of the study population are presented in Table 1. To protect patient privacy, cell counts of 10 or fewer were suppressed.

**Table 1.** Demographic and Clinical Characteristics of Patients with Neurocysticercosis and Comparison Patients in the UC Health System.

| Characteristics | NCC<br>n (%) | Non-NCC<br>n (%) | Test<br>Statistic | df | Effect Size<br>(95% CI,<br>where<br>applicable)<sup>‡</sup> | p-value |
| --- | --- | --- | --- | --- | --- | --- |
| Age (years),<br>mean (SD) | 57.4 (17.0) | 48.6 (23.6) | t = -12.7 | 1010.9 | -8.84<br>(-10.21, -7.48) | < 0.001**a |
| Sex |  |  | - | - | 0 | 0.460<sup>b</sup> |
| Male | 307 (43.1) | 3234 (45.4) |  |  |  |  |
| Female | 406 (56.9) | 3885 (54.5) |  |  |  |  |
| Other | 0 | ≤ 10 |  |  |  |  |
| Unknown | 0 | ≤ 10 |  |  |  |  |
| Race |  |  | - | - | 0.363 | < 0.001**b |
| Asian | 31 (4.35) | 700 (9.82) |  |  |  |  |
| Black or African American | ≤ 10 | 340 (4.77) |  |  |  |  |
| Hispanic or Latino | 534 (74.9) | 1445 (20.27) |  |  |  |  |
| Multirace | ≤ 10 | 211 (2.96) |  |  |  |  |
| Native Hawaiian or Other Pacific Islander | ≤ 10 | 25 (0.35) |  |  |  |  |
| White | 59 (8.27) | 2928 (41.1) |  |  |  |  |
| Other Race | 28 (3.93) | 481 (6.75) |  |  |  |  |
| Unknown | 46 (6.45) | 1000 (14.0) |  |  |  |  |
| **Ethnicity** | | | $\chi^2 = 1032.6$ | 2 | 0.363 | < 0.001**c |
| Hispanic or Latino | 534 (74.9) | 1445 (20.3) |  |  |  |  |
| Not Hispanic or Latino | 118 (16.5) | 4538 (63.6) |  |  |  |  |
| Unknown | 61 (8.56) | 1147 (16.1) |  |  |  |  |
| **Primary Spoken Language** | | | $\chi^2 = 1398.3$ | 3 | 0.422 | < 0.001**c |
| English | 237 (33.2) | 5490 (77.0) |  |  |  |  |
| Spanish | 335 (47.0) | 385 (5.40) |  |  |  |  |
| Other | 11 (1.54) | 186 (2.61) |  |  |  |  |
| Unknown | 130 (18.2) | 1069 (15.0) |  |  |  |  |
| **Primary Insurance** |  |  | - | - | 0.196 | < 0.001**b |
| Medicare | 136 (19.1) | 1017 (14.3) |  |  |  |  |
| Medicaid | 342 (48.0) | 1532 (21.5) |  |  |  |  |
| Department of<br>Veteran's<br>Affairs | ≤ 10 | 117 (1.64) |  |  |  |  |
| Private<br>Insurance | 205 (28.8) | 3752 (52.6) |  |  |  |  |
| Unknown | 27 (3.79) | 712 (9.99) |  |  |  |  |
| **Area**<br>**Deprivation**<br>**Index (ADI)** | 5.89 ± 2.6<sup>†</sup> | 4.73 ± 2.8<sup>†</sup> | t = 11.12 | 864.21 | 1.16<br>(0.95, 1.36) | < 0.001**a |
| **Mortality** |  |  |  |  |  |  |
| N deceased (%) | 61 (8.56) | 493 (6.91) | $\chi^2 = 2.41$ | 1 | 0.01 | 0.12<sup>d</sup> |
| Avg Age at<br>Death | 64.6 | 68.5 | t = -1.88 | 79.76 | -3.90<br>(-8.03, 0.22) | 0.063<sup>a</sup> |

\* $p < 0.05$ \*\* $p < 0.001$
a Welch Two Sample T-Test
b Fisher's exact test with Monte Carlo approximation (100,000 simulations)
c Pearson's Chi-square test
d Pearson's Chi-square test with Yates' continuity correction
<sup>†</sup> Area Deprivation Index (ADI) was missing for 19 patients in the NCC group and 395 patients in the comparison group.
<sup>‡</sup> Effect sizes are reported as mean differences for continuous variables and Cramér's V for categorical variables. 95% confidence intervals are reported for mean differences.

Patients with NCC were older on average than patients in the comparison group (57.4 years and 48.6 years, respectively; *p* < 0.0001). Sex distribution did not differ significantly across groups (*p* = 0.460).

Race, ethnicity, and primary spoken language differed significantly across groups (*p* < 0.0001 for all). Patients with NCC were disproportionately Hispanic or Latino and Spanish- speaking relative to the comparison group, similar to the demographic profile of populations from some NCC-endemic regions. Spanish was the most common primary language among NCC patients (n = 336, 47.1%) compared with the comparison group (n = 388, 5.44%), while English was the predominant language among patients without NCC (n = 5523, 77.5%).

Insurance coverage and neighborhood socioeconomic disadvantage also differed significantly across groups (*p* < 0.0001 for both). Medicaid was the most common insurance type among NCC patients, while private insurance was more common among the comparison group. Mean ADI scores were higher among NCC patients compared with the comparison group, indicating residence in more socioeconomically disadvantaged neighborhoods.

The NCC group had a slightly higher proportion of deceased patients compared to the comparison group (8.56% vs 6.91%), however this difference was not significant (*p* = 0.12). The NCC group also had a slightly lower average age at death compared to the comparison (64.6 vs 68.5), however this difference was not statistically significant (*p* = 0.063).

### Neurologic Comorbidities

Neurologic and psychiatric comorbidities were significantly more common among patients with NCC relative to the comparison group across nearly all conditions examined. Descriptive statistics and tests of association are presented in Table 2, and univariate logistic regression results are presented in Table 3 and Fig 1.

**Fig 1.**
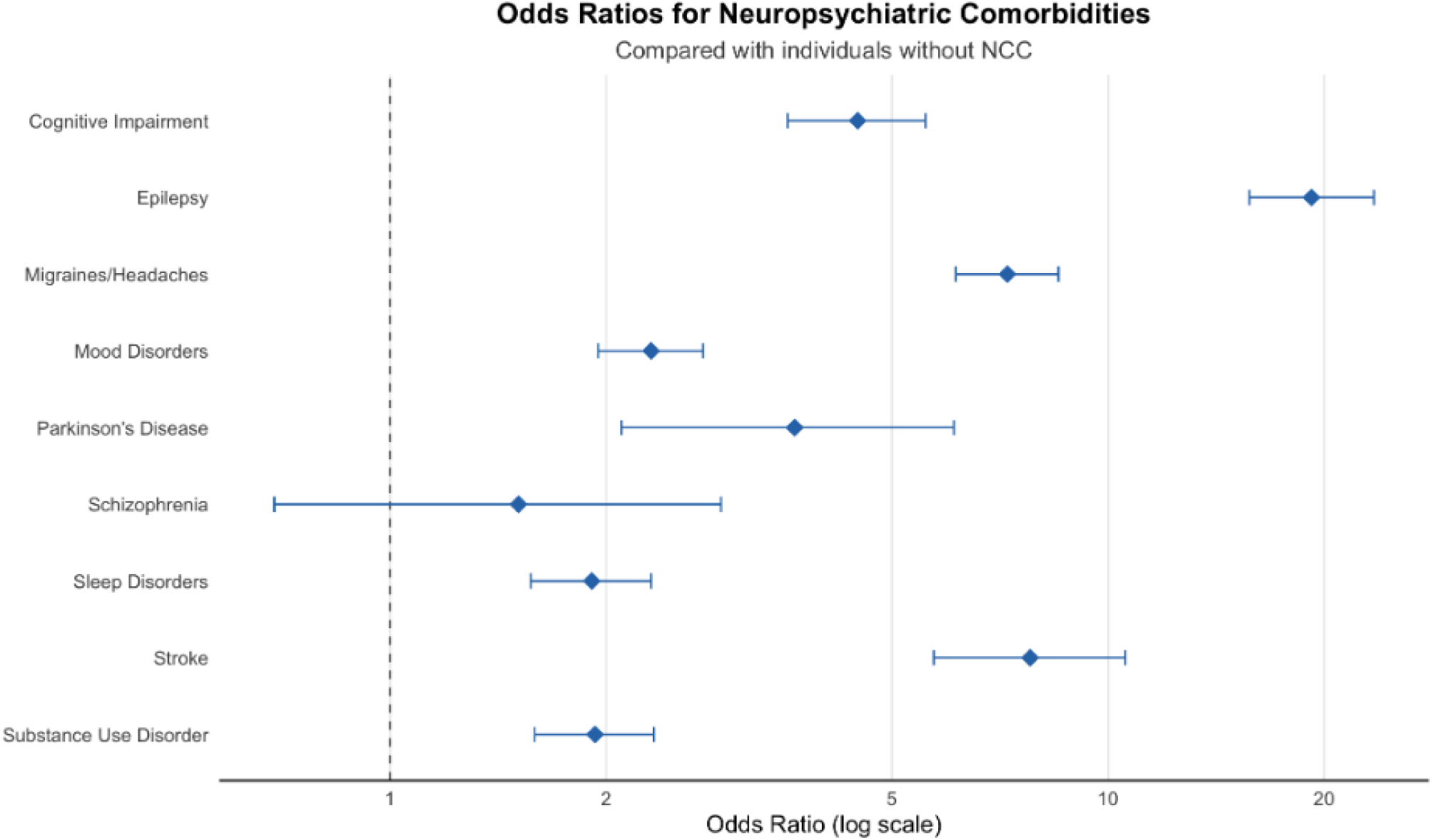
Odds Ratios for Neuropsychiatric Comorbidities Among People with NCC Compared with Those Without NCC.

**Table 2.** Neurologic Comorbidities Among Patients with Neurocysticercosis in the UC Health System.

| Comorbidity | NCC<br>n (%) | Non-NCC<br>n (%) | Test<br>Statistic<br>(chi) | df | Effect Size<sup>‡</sup> | p-value |
| --- | --- | --- | --- | --- | --- | --- |
| Migraines and Headaches | 334 (46.8) | 774 (10.9) | 689.1 | 1 | 0.297 | <0.0001**a |
| Cognitive Impairment | 128 (18.0) | 332 (4.66) | 205.14 | 1 | 0.162 | <0.0001**a |
| Mood Disorders | 234 (32.8) | 1243 (17.4) | 99.38 | 1 | 0.113 | <0.0001**a |
| Parkinson's Disease | 19 (2.66) | 53 (0.74) | - | - | 0.057 | <0.0001**b |
| Schizophrenia | <10 | 60 (0.84) | - | - | 0.006 | 0.287<sup>b</sup> |
| Substance Use Disorders | 156 (21.9) | 903 (12.7) | 46.3 | 1 | 0.077 | <0.0001**<sup>a</sup> |
| Stroke | 75 (10.5) | 106 (1.49) | 230.6 | 1 | 0.173 | <0.0001**<sup>a</sup> |
| Sleep Disorders | 154 (21.6) | 899 (12.6) | 44.3 | 1 | 0.075 | <0.0001**<sup>a</sup> |

\* $p < 0.05$ \*\* $p < 0.001$
<sup>a</sup> Pearson's Chi-squared test with Yates' continuity correction
<sup>b</sup> Fisher's exact test
<sup>‡</sup> Effect sizes are reported as Cramér's V for categorical variables.

**Table 3.** Logistic Regression Analysis of Neurologic Comorbidities in Patients with Neurocysticercosis Compared with Patients Without Infection.

| Comorbidity | Odds Ratio (95% CI) | *p*-value |
| --- | --- | --- |
| Cognitive Impairment | 4.48 (3.58, 5.57) | <0.0001** |
| Epilepsy | 19.20 (15.74, 23.46) | <0.0001* |
| Migraines or Headaches | 7.24 (6.14, 8.53) | <0.0001** |
| Mood Disorders | 2.31 (1.95, 2.73) | < 0.0001** |
| Parkinson's Disease | 3.66 (2.10, 6.10) | <0.0001** |
| Schizophrenia | 1.51 (0.69, 2.89) | 0.255 |
| Sleep Disorders | 1.91 (1.57, 2.31) | < 0.0001** |
| Stroke | 7.79 (5.72, 10.57) | < 0.0001** |
| Substance Use Disorders | 1.93 (1.59, 2.33) | < 0.0001** |

\* $p < 0.05$ \*\* $p < 0.001$
CI = Confidence Interval

Following epilepsy, migraines and headaches showed the strongest associations with NCC among all comorbidities examined. Relative to the comparison group, NCC was associated with 7.24 times higher odds (95% CI 6.14–8.53) of having migraines and headaches. Cognitive impairment and stroke were also strongly associated with NCC, with odds ratios (OR) of 4.48 (95% CI 3.58, 5.57) for cognitive impairment and 7.79 (95% CI 5.72, 10.57) for stroke. Patients with NCC also were associated with 3.66 times the odds of having Parkinson’s disease (95% CI 2.10-6.10) compared to the comparison group. NCC was associated with elevated odds of having mood disorders (OR 2.31, 95% CI 1.95–2.73), compared to the comparison group. NCC patients were also associated with 93% higher odds of having substance use disorders (95% CI 1.59, 2.33) and 91% higher odds of having sleep disorders (95% CI 1.57, 2.31). NCC was not significantly associated with an increased odds of schizophrenia (*p* = .255).

### Epilepsy and Associated Factors

In unadjusted logistic regression analyses (Table 4), NCC was strongly associated with epilepsy compared to patients without NCC (*p* < .001). Patients with NCC had significantly higher odds of epilepsy than the comparison group (OR 19.20, 95% CI 15.74–23.46).

**Table 4.** Unadjusted Logistic Regression Model Evaluating the Association Between Neurocysticercosis and Epilepsy (n = 7843)

|  | Odds Ratio (95% CI) | p-value |
| --- | --- | --- |
| *NCC Diagnosis Status* |  |  |
| None | 1 [Reference] |  |
| Neurocysticercosis | 19.20 (15.74-23.46) | < 0.0001* |

\* $p < 0.05$ \*\* $p < 0.001$
CI = Confidence Interval

After adjusting for age, sex, ethnicity, and ADI (Table 5), these associations remained substantial and largely unchanged. NCC was associated with 20.08 times the odds of epilepsy (95% CI 15.62–25.93) in the adjusted model. Among the covariates, male sex was associated with 26% higher odds of epilepsy (OR 1.26, 95% CI 1.03–1.55, *p* = 0.024) and increasing age with slightly lower odds (OR 0.99, 95% CI 0.99–1.00, *p* = 0.019), though the magnitude of this association was minimal. Ethnicity and ADI were not significantly associated with epilepsy in the adjusted model (*p* = 0.584 and *p* = 0.097, respectively).

**Table 5.**
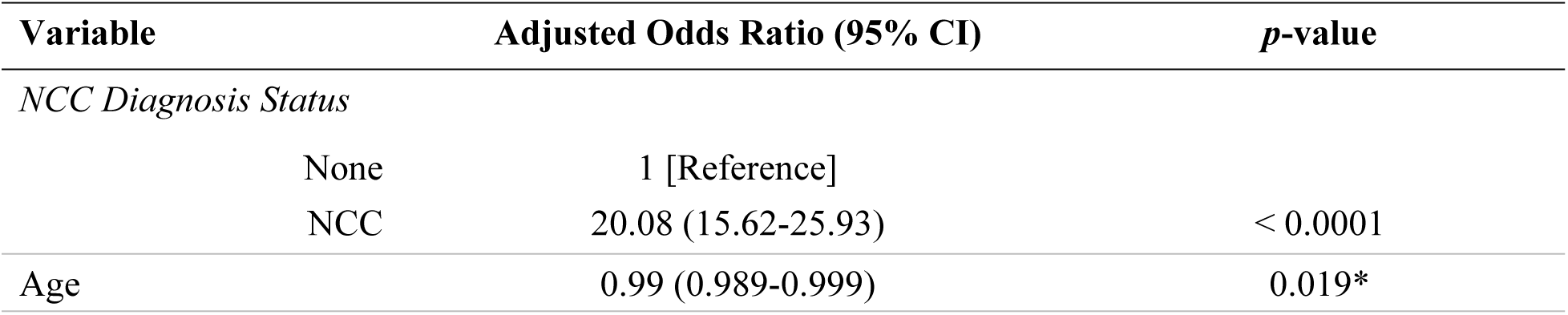
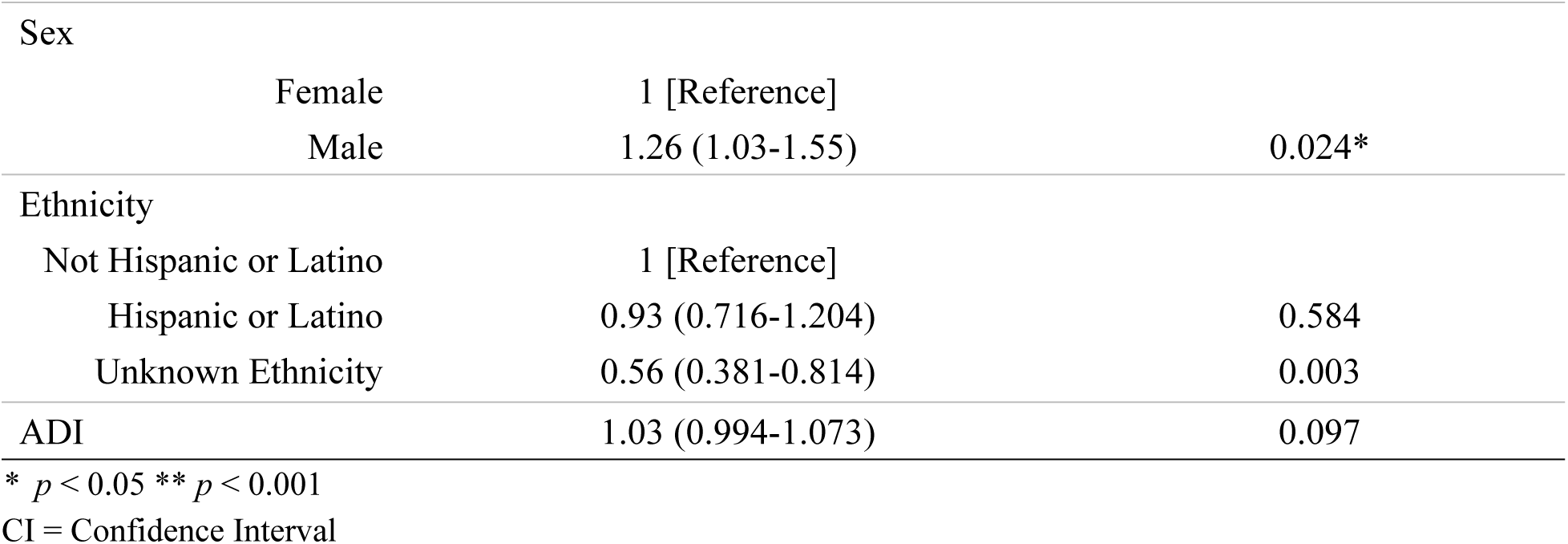
Adjusted Logistic Regression Model Evaluating Factors Associated with Epilepsy (n = 7422)

## Discussion

In this large, multi-center study, we identified 713 patients with NCC within the UC Health system using electronic health record data spanning a 14-year period (2012–2026), corresponding to a prevalence of 6.17 cases per 100,000 patients. Although the UC system is not fully representative of the California population, its large, statewide network and diverse patient population provide an important window into the burden of NCC in the state. The prevalence observed in this population demonstrates that NCC remains an important cause of neurologic disease in California. This finding is particularly noteworthy because the observed prevalence should not be interpreted as a population-based estimate and likely underestimates the true burden of disease. Populations at higher risk for NCC, including Hispanic immigrants and individuals living in rural areas, often face barriers to healthcare access, diagnosis, and specialty referral,[11,12] likely limiting their representation within an academic health system. Thus, the burden of NCC observed in the UC system may represent only a fraction of the disease burden across California.

Importantly, the individuals with NCC identified in this study had multiple demographic and socioeconomic indicators of social vulnerability. Compared with the overall UC Health population, patients with NCC were older and disproportionately identified as Hispanic or Latino, were more likely to report Spanish as their primary language, were more frequently insured through Medicaid, and resided in neighborhoods with greater socioeconomic disadvantage as measured by the ADI.

Immigrant populations in the United States may encounter challenges including limited insurance coverage, language barriers, high out-of-pocket costs for diagnostic testing, and reduced access to preventive and specialty care, all of which may delay recognition and treatment of neurologic disease.[11,12] Efforts to improve access to culturally and linguistically appropriate care, timely neurologic evaluation, and specialty referral for populations at increased risk may help reduce delays in diagnosis, mitigate neurologic morbidity, and advance health equity in California.

In addition to the social and structural vulnerabilities observed in this population, our findings demonstrate the substantial neurologic and psychiatric morbidity associated with NCC. Patients with NCC had significantly higher odds of epilepsy, headaches, cognitive impairment, mood disorders, stroke, substance use disorders, sleep disorders, and Parkinson’s disease compared with the overall UC Health population. The strong association between NCC and epilepsy is consistent with its established role as one of the leading causes of acquired epilepsy worldwide.[1,3,5] Similarly, the increased prevalence of headaches and cognitive impairment aligns with prior studies identifying these as among the most common neurologic manifestations of NCC.[1–3,5] In contrast, the associations observed with mood disorders, schizophrenia, sleep disorders, and substance use disorders have received comparatively less attention in the literature,[5] suggesting that the burden of psychiatric and neurologic comorbidity associated with NCC may be broader than has been traditionally recognized.

Although the clinical manifestations of NCC have been well described in *T. solium* endemic regions of Latin America, Africa, and Asia, large-scale epidemiologic studies in the United States remain limited.[1–4,5,6] The neurologic and psychiatric comorbidities identified in this population are broadly consistent with observations from endemic settings, indicating that the clinical burden of NCC extends to United States healthcare systems. By characterizing these associations within California’s largest integrated academic healthcare network, our study provides contemporary evidence that the impact of NCC in the United States extends beyond epilepsy alone and encompasses a broad spectrum of neurologic and psychiatric disease. The high burden of neurologic and psychiatric comorbidities likely compounds the existing social and structural vulnerabilities experienced by this population.

The elevated burden of neurologic and psychiatric comorbidity observed in this cohort underscores the substantial clinical needs of individuals affected by NCC. Many patients experienced multiple neurologic comorbidities, reflecting the complex, multisystem nature of the disease, and the need for coordinated neurologic care. These clinical challenges are likely to further compound the social and structural vulnerabilities identified in this population, as conditions such as epilepsy may adversely affect cognition, mood, functional independence, educational attainment, employment opportunities, and overall quality of life.[21]

Epilepsy, the comorbidity most strongly associated with NCC in this study, is itself associated with substantial long-term morbidity. The economic, psychosocial, physical, and mental burden of epilepsy is substantial for affected individuals, their families, and health systems, and the condition increases the risk of premature death up to three times compared with the general population.[10]

Although our study did not directly evaluate diagnostic or treatment delays, the demographic and socioeconomic characteristics of this cohort suggest that barriers to healthcare access may place many patients at increased risk for delayed recognition and management of both NCC and its neurologic sequelae. Newly diagnosed NCC has been associated with mild cognitive deficits and significant reductions in quality of life, particularly in social and mental function, which improve following treatment.[22] These findings highlight the importance of improving timely recognition of NCC and facilitating access to comprehensive neurologic care, particularly for individuals who have immigrated from or traveled to *T. solium* endemic regions.

NCC was strongly associated with epilepsy (aOR 20.08, 95% CI 15.62–25.93), but this association remained largely unchanged after adjusting for age, sex, ethnicity, and ADI. This suggests that the relationship between NCC and epilepsy is not substantially explained by these demographic or socioeconomic factors. Male sex was associated with 26% increased odds of epilepsy, and older age was associated with slightly lower odds of epilepsy. Hispanic or Latino ethnicity and ADI were not significantly associated with epilepsy after adjustment; however, patients with unknown ethnicity had significantly lower odds of epilepsy. This likely reflects poorer quality in documentation in the EHR rather than a true epidemiologic relationship and should therefore be interpreted with caution.

These findings should be interpreted with the cross-sectional design of this study in mind. Because NCC and epilepsy diagnoses were not temporally ordered, directionality and causality cannot be established. It is possible that NCC precedes and contributes to epilepsy, but it is also possible that patients presenting with seizures were more likely to undergo neuroimaging that led to NCC detection, which could inflate the observed association. Longitudinal data linking the timing of NCC incidence to epilepsy onset are needed to clarify this relationship.

This study has several notable strengths. The use of a large dataset encompassing six University of California health systems allowed for the identification of a relatively large population of individuals with NCC. Given the relative rarity of NCC, or at least NCC diagnoses, in the U.S., this scale provides valuable insight into disease patterns in a non-endemic setting for *T. solium*. This approach also allowed for a highly cost-effective analysis of a geographically and demographically diverse patient population. This study examined a broader range of neurologic comorbidities than many prior NCC studies, providing a comprehensive assessment of the burden of neurologic complications associated with the disease.[5]

Several limitations should be considered when interpreting these findings. First, as mentioned previously, the cross-sectional design does not allow for the assessment of temporal relationships, limiting causal inference between NCC and observed neurologic outcomes. The analysis relied on diagnostic coding within the EHR, which may be subject to variable degrees of undercoding or misclassification. NCC, in particular, is difficult to diagnose based only on symptoms, requiring costly neuroimaging and often serologic testing.[1–3,7] We omitted cases with only a documented diagnosis of cysticercosis or *T. solium* infection, some of which may have been true NCC cases. This potential underestimation of NCC prevalence may bias measures of association reported in this study.[23] Neuroimaging data were not available to further investigate these cases. It is known that variables such as lesion location, size, and number impact the clinical presentation of NCC,[1] however, without further information, we were not able to assess how these factors influenced clinical outcomes. It is also likely that the study population is biased towards sicker cases, since patients with more severe disease may have more points of contact with the health system, with more opportunity for providers to input an accurate diagnosis code. In contrast, more mild or minimally symptomatic cases are more likely to go undiagnosed. Potentially important variables such as immigration or travel history were not available in the dataset. It is also likely that some individuals with NCC remain undiagnosed due to limitations in health care access. Since the comparison group included randomized patients, they may have interacted less with the health system, which could account for the lower rates of some comorbidities.

Our findings demonstrate that NCC remains an important but underrecognized neurologic disease in California, disproportionately affecting socially and medically vulnerable populations while carrying a substantial burden of neurologic and psychiatric comorbidity. Because NCC may remain asymptomatic for years, clinically recognized cases likely represent only a fraction of the true disease burden. Improving provider awareness, maintaining a high index of suspicion among patients with epidemiologic risk factors, and facilitating timely diagnostic evaluation may enable earlier recognition and treatment, with the potential to reduce long-term neurologic morbidity and reduce health disparities. Clinicians should consider NCC in the differential diagnosis of individuals who have immigrated from or traveled to *T. solium* endemic regions presenting with seizures or other unexplained neurologic or psychiatric manifestations. As migration from endemic regions continues and patients with NCC seek care throughout the United States, improving recognition of this disease will be essential to reducing preventable neurologic disability and advancing health equity.

Future studies using longitudinal data are needed to better characterize the temporal relationships between NCC and its associated neurologic outcomes, particularly epilepsy and cognitive impairment, and to identify clinical predictors of long-term neurologic morbidity.

## Acknowledgements

The authors thank the Center for Data-driven Insights and Innovation at UC Health (CDI2; https://www.ucop.edu/uc-health/departments/center-for-data-driven-insights-and-innovations-cdi2.html), for its analytical and technical support related to use of the UC Health Data

Warehouse and related data assets. The authors would also like to thank the Enterprise Analytics and Data Services (EADS) team at UC Davis Health for their support in obtaining access to the necessary databases and reviewing the SQL code developed by the authors for this study. The EADS team is supported by the National Center for Advancing Translational Sciences, National Institutes of Health, through grant number UL1 TR001860. The content is solely the responsibility of the authors and does not necessarily represent the official views of the NIH.

## Data Availability Statement

Data that support the findings of this study were obtained from the University of California Health Data Warehouse (UCHDW)/University of California Data Discovery Platform (UCDDP), a HIPAA limited data set which contains patient data from all 6 UC Health academic medical centers (Davis, Irvine, Los Angeles, Riverside, San Diego, and San Francisco). Due to privacy and confidentiality restrictions, the data are not publicly available. Minimal de-identified aggregate data in the form of tables may be available from the corresponding author on reasonable request and subject to corresponding author’s institutional approval.

The code used to perform the analyses in this study is currently maintained in a private repository and will be made publicly available without restriction upon publication of this article.

## Table of Abbreviations

NCC: Neurocysticercosis
UC: University of California
CNS: Central Nervous System
WHO: World Health Organization
OMOP: Observational Medical Outcomes Partnership
HIPAA: Health Insurance Portability and Accountability Act
UCHDW: University of California Health Data Warehouse
EHR: Electronic Health Record
CDI2: Center for Data-driven Insights and Innovation
OHDSI: Observational Health Data Sciences and Informatics
ICD: International Classification of Diseases
SNOMED: Systematized Nomenclature of Medicine
ADI: Area Deprivation Index
OR/ORs: Odds Ratio(s)
CI/CIs: Confidence Interval(s)

## Author Contributions

Tayf Mohammed Islam: Data Curation, Methodology, Formal Analysis, Methodology, Writing – Original Draft Preparation

Samantha Eve Allen: Conceptualization, Methodology, Project Administration, Supervision,

Writing – Review & Editing

